# Excitatory inhibitory imbalance underlies hippocampal atrophy in individuals with 22q11.2 Deletion Syndrome with psychotic symptoms

**DOI:** 10.1101/2022.09.06.22279580

**Authors:** Valentina Mancini, Muhammad G. Saleh, Farnaz Delavari, Joëlle Bagautdinova, Stephan Eliez

## Abstract

**Background:** Abnormal neurotransmitter levels have been reported in subjects at high risk for schizophrenia, leading to a shift in the excitatory/inhibitory balance. However, it is unclear if these alterations are predating the onset of clinically relevant symptoms. Our aim was to explore in vivo measures of excitatory/inhibitory balance in 22q11.2 deletion carriers, a population at high genetic risk for psychosis.

**Methods:** Glx (glutamate + glutamine) and GABA+ concentrations were estimated in the anterior cingulate cortex (ACC), superior temporal gyrus (STG) and hippocampus using a MEGAPRESS sequence and the Gannet toolbox in 52 deletion carriers and 42 controls. T1-weighted images were acquired longitudinally and processed with Freesurfer v.6.0 to extract hippocampal volume. Subgroup analyses were conducted in deletion carriers with psychotic symptoms identified by means of SIPS.

**Results:** While no differences were found in the ACC, deletion carriers had higher levels of Glx in the hippocampus and STG, and lower levels of GABA+ in the hippocampus compared to controls. We additionally found a higher Glx concentration in the hippocampus of psychotic compared to non-psychotic deletion carriers. Finally, more pronounced hippocampal atrophy and increased functional variability were both significantly associated with increased Glx levels in deletion carriers.

**Conclusions:** This study provides evidence for an excitatory/inhibitory imbalance in temporal brain structures of deletion carriers, with a further hippocampal Glx increase in individuals with psychotic symptoms that was associated with hippocampal atrophy and abnormal function. These results support theories proposing abnormally enhanced glutamatergic neural transmission as a mechanistic explanation for hippocampal atrophy via excitotoxicity. Overall, our results highlight a central role of glutamate in the hippocampus of individuals at genetic risk for schizophrenia.

## Introduction

While traditional accounts of schizophrenia pathophysiology pointed to a major dysregulation of the dopaminergic system, abnormalities in the glutamatergic and gamma-aminobutyric acid (GABA)-ergic systems have become increasingly acknowledged (1). Glutamatergic dysfunction is believed to stem from the hypofunction of N-methyl-D-aspartate receptors (NMDAR) expressed on parvalbumin positive interneurons (PVI) (2). This decreased excitatory drive onto PVI indirectly triggers the disinhibition of pyramidal neurons, thereby increasing their glutamate release (3). This model has been validated by histological data showing decreased NMDAR binding in medial temporal lobe post-mortem samples from patients with schizophrenia (4) and clinical data indicating that the administration of NMDAR antagonists, such as ketamine (5, 6), and autoimmune encephalitis with autoantibodies targeting NMDAR (7, 8) can induce psychotic symptoms in previously healthy individuals. Genetic evidence further supports this hypothesis, as a schizophrenia genome-wide association study highlighted the involvement of several genes implicated in glutamatergic neural transmission (9).

One of the main techniques to estimate the concentration of glutamate and GABA levels *in vivo* in humans is proton magnetic resonance spectroscopy (MRS). Meta-analytic MRS studies reported increased glutamatergic concentration across several brain regions in patients with schizophrenia (10) and a correlation between higher brain glutamate and illness severity (11). More recently, advances in neuroimaging have permitted the estimation of GABA concentrations in the brain. Studies thus far have reported mixed findings, including decreased or normal GABA levels (12–15) and an association between GABA levels and cognition in patients with schizophrenia (15). Conflicting results were similarly found for individuals at clinical-risk for (CHR) or at the first episode of psychosis (FEP), with some studies reporting increased glutamate concentration (16, 17) whereas others reporting no differences compared to healthy controls (13, 18). Finally, evidence from genetic models of schizophrenia such as the copy number variant 22q11.2 deletion syndrome (22q11DS) have also yielded mixed evidence (19–21).

While differences in methodology and regions of interest exist, age likely plays a key role in the heterogeneity of prior findings. During late adolescence, many dramatic changes in the balance between excitatory and inhibitory neural transmission are observed, including an increase of inhibitory and a decrease of excitatory synapses (22–24). The maturation of inhibitory GABAergic results in a progressive reduction of the E/I ratio during adolescence (25). Given these dynamic changes in the E/I balance, it becomes critical to examine neurotransmitter levels in a developmental context to accurately delineate potential alterations.

Genetically defined syndromes, such as 22q11DS, represent a unique opportunity for developmental studies, as their early postnatal diagnosis offers the possibility to follow-up with affected individuals over time. Deletion carriers have a strong predisposition for neuropsychiatric illnesses, including anxiety, attention deficit hyperactivity disorder (ADHD), obsessive compulsive disorder (OCD) and mood disorders, with an estimated 40% lifetime risk to develop a psychotic disorders by adulthood (26). 22q11DS has received increased attention as a high-penetrance model for schizophrenia due to the presence of critical genes for cortical circuits formation and neurotransmission - including glutamatergic, GABAergic and dopaminergic signaling - in the deleted region (27, 28). Given the important remodeling events occurring in these neurotransmitters levels during development (22, 23, 25), it thus becomes likely that genetic insults affecting relevant genes may lead to an imbalance in excitatory and inhibitory transmission in late maturing brain regions (22, 23). In support of this hypothesis, studies in the homologous mice model of 22q11DS have shown abnormally enhanced glutamate release resulting from altered calcium kinetics (29) and hypofunction of PVI (30) in adult animals. However, the E/I balance has thus far not been developmentally investigated in human 22q11 deletion carriers.

In this study, we leveraged advances in MRS sequences permitting the estimation of GABA concentrations to measure excitatory and inhibitory neurotransmitters levels in deletion carriers versus controls. For our MRS analysis, we selected three late-maturing brain regions that over the years have emerged as being functionally and structurally affected in 22q11 deletion carriers and idiopathic psychosis: anterior cingulate cortex (ACC) (31–34), superior temporal cortex (STC) (34– 40) and hippocampus (41–44). In particular, there is evidence of altered maturational trajectories of the STC and hippocampus (37, 42) as well as disrupted functional connectivity (43) in deletion carriers experiencing psychotic symptoms, highlighting the importance of investigating markers of E/I balance in these regions during critical developmental periods. Based on previous studies (11, 14, 20, 45), we expected to find higher levels of glutamate and lower levels of GABA in deletion carriers, with a more pronounced increase of glutamate in individuals experiencing psychotic symptoms. Additionally, we conducted exploratory analyses to determine whether neurometabolite levels were associated with age, known hippocampal atrophy (42), and abnormal hippocampal functional activity (43). Finally, we examined age and age-by-metabolite interactions in the hippocampus of deletion carriers with or without psychosis symptoms.

## Materials and Methods

### Participants

Individuals without (healthy controls, HC) and with 22q11DS were recruited from the longitudinal 22q11DS Swiss Cohort. Recruitment was carried out through word of mouth, community announcements, and advertisements aimed at parents’ associations. At baseline and follow-up visits, individuals with 22q11DS and HC underwent magnetic resonance imaging (MRI) and clinical assessment. Inclusion criteria for the present study were age range 7 and 30 years old and the presence of a 22q11.2 microdeletion confirmed by quantitative fluorescent polymerase chain reaction (QF-PCR) for patients. Exclusion criteria for controls were any past or present neurological or psychiatric disease, use of psychotropic medications, psychopathology, learning difficulties or prematurity. The occurrence of attenuated psychotic symptoms was assessed in deletion carriers by means of the Structured Interview for Psychosis-risk Syndromes (SIPS) (46). Deletion carriers were divided in subgroups according to the presence of moderate-to-severe psychotic symptoms, using a cut-off score of 3 or higher in at least one of the corresponding items for positive symptoms of the SIPS. Written informed consent was obtained from participants and/or their parents. The study was approved by the cantonal ethics committee of Geneva for research and conducted according to the Declaration of Helsinki.

### Data Acquisition

Brain MRI and MRS experiments were conducted on 3T MRI scanner (Siemens MAGNETOM Prisma, Erlangen, Germany) using a 20-channel head coil. A T1-weighted images with a three-dimensional volumetric pulse were acquired from each subject using the following parameters: TR/TE□2500/3□ms; flip angle□8°; acquisition matrix□256□×□256; field of view□235□mm; slice thickness□3.2□mm; and 192 slices. After T1 images, resting state fMRI data were recorded with a sequence of 8 minutes (voxel size 1.84×1.84×3.2 mm, 38 slices, TR/TE 2400/30 ms, flip angle 85°). During the resting-state session, participants were instructed to focus on a cross displayed on the computer screen. The T1-images were used to guide proton (1H) MRS voxel positioning in the anterior cingulate cortex (ACC), left hippocampus, and left superior temporal cortex (STC), as shown in Figure 1. Subjects were scanned using the Mescher-Garwood point-resolved spectroscopy (MEGA-PRESS) Siemens prototype sequence (47, 48) for the measurement of GABA with macromolecules and homocarnosine (GABA+) at 3.0 ppm with the following parameters: TR/TE 1500/68 ms; spectral width: 2000 Hz; editing pulse offset (ON/OFF) 1.9/7.5 ppm; bandwidth of the editing pulse specified on the scanner 60 Hz; 2048 data points; and voxel sizes and number of transients 15×35×40 mm^3^ and 196 for ACC, 20 × 30 × 40 mm^3^ and 300 for left hippocampus, and 30×30×30 mm^3^ and 240 for STC. Finally, unsuppressed water acquisitions were performed with the same region-specific parameters, except 16 transients for concentration quantification.

**Figure 1.**
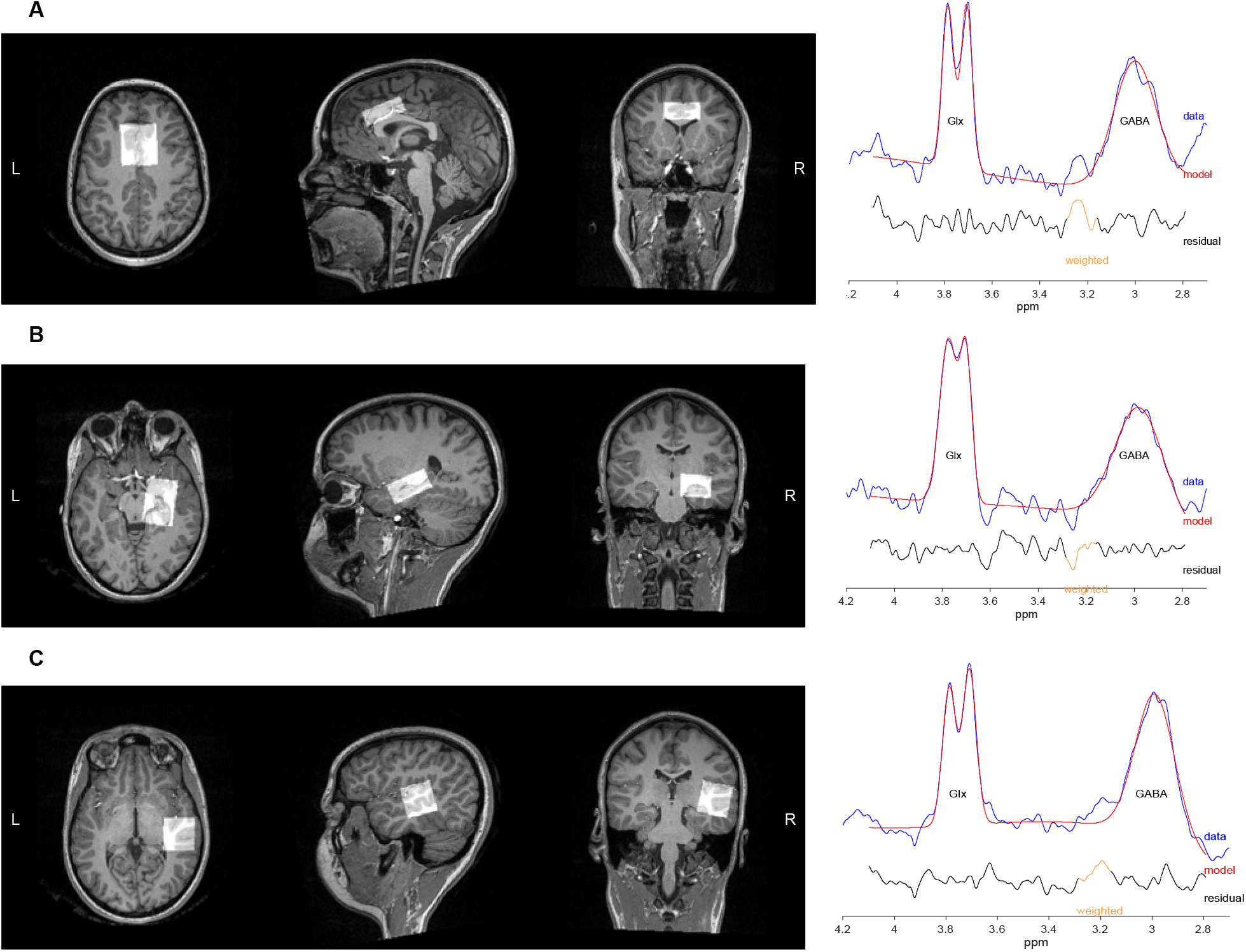
Voxel positions for the MEGAPRESS sequence and sample spectra. Voxel positions for: **A)** Anterior cingulate cortex (ACC) placed on the midline above the genu of the corpus callosum, with the anterior edge of the ROI never exciding the genu of the corpus callosum, as shown in Fig 1a (voxel: 15×35×40; averages: 196); **B)** left hippocampus: placed around the hippocampus including head, the body and part of the tail, as shown in Fig 1b (voxel: 20×30×40; averages: 300); **C)** left superior temporal cortex (STC): placed at the level of the left superior temporal gyrus, comprising the Heschl’s gyrus, as shown in Fig 1c (voxel: 30×30×30; averages: 240). On the right side the corresponding spectra from one participant. Glx= composite signal of glutamate and glutamine; GABA= gamma-aminobutyric acid + macromolecules; L= left; ppm= parts per million; R= right.

### MRI analyses

T1-weighted images underwent fully automated image processing with FreeSurfer version 6 (49), comprising skull stripping, intensity normalization, reconstruction of the internal and external cortical surface and parcellation of subcortical brain regions (50). Manual quality control was performed for every MRI scan. Longitudinal T1-weighted images were available for 52 deletion carriers, and we extracted the hippocampal volume with FreeSurfer (51) from images acquired at the same time as the spectroscopy (H1-MRS_V2_) and at the previous visit on average 3.14 years before (H1-MRS_V1_). A percentage score of atrophy was calculated as follows: 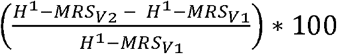 and used for following statistical analyses.

### Rs-fMRI preprocessing and analyses

fMRI preprocessing was performed using an adapted and previously published pipeline (52) using Statistical Parametric Mapping (SPM12, Wellcome Trust Centre for Neuroimaging, London, UK: http://www.fil.ion.ucl.ac.uk/spm/) and functions of the Data Processing Assistant for Resting-State fMRI (DPARSF; (53)) and Individual Brain Atlases using Statistical Parametric Mapping (IBASPM; (54)) toolboxes. After realignment of functional scans, we applied spatial smoothing with an isotropic Gaussian kernel of 6 mm full width half maximum (FWHM) and co-registered structural scans to the functional mean. Structural images were segmented with the SPM12 Segmentation algorithm (55) and a study-specific template was generated using Diffeomorphic Anatomical Registration Through Exponentiated Lie algebra (DARTEL; (56)). Then, the first 10 functional scans were excluded from the analysis, mean white-matter and CSF signals were regressed from the BOLD time series, which were then filtered with a bandwidth of 0.01 Hz to 0.1 Hz. For a more extended correction of motion artifacts, we further applied motion scrubbing (57), excluding frames with a framewise displacement of more than 0.5 mm, as well as one frame before and two frames after. A mask of hippocampus as region of the interest was selected for the left hippocampus based on the atlas Automated Anatomical Labeling (AAL) (58). Using the study specific DARTEL template the masks were spatially transformed into the native space. Voxel-wise BOLD signal variability (BSV) was calculated as the standard deviation the voxel’s time series in the native subject space (52). This resulted in a voxel-wise variability map in native space. The regional BSV for the left hippocampus was calculated as the average of voxel-wise BSV values of voxels within that region.

### MRS analyses

All spectra data were processed using Gannet (version 3.1.5) (59). After frequency and phase correction (FPC) of transients, data were combined to generate GABA-edited spectra. The GABA+, glutamate and glutamine (Glx), and the unsuppressed water signals were modelled to calculate GABA and Glx concentrations in institutional units (IU). T1-weighted images were segmented using SPM12 (55) to calculate gray matter, white matter, and CSF voxel tissue fractions in the ^1^H-MRS voxels. Subsequently, metabolite concentrations were corrected for partial volume effects using the tissue fractions and relaxation effects with the Gasparovic method (60). Data quality was assessed using the Cr signal at 3 ppm to estimate main magnetic field (B_0_) drift (before FPC) and using the unsuppressed water signal linewidth at full width at half maximum. The GABA and Glx fit errors generated by Gannet (defined as the ratio of the SD of the fit residual to the amplitude of the modeled peak) were used for assessing modeling errors. As in previous papers (61), data with fit errors exceeding 2 standard deviation above the mean of all the data (15%) were excluded from further analyses.

### Statistical analyses

Statistical analyses were conducted in Matlab 2018b (Mathworks) and R studio (http://www.rstudio.com/). Shapiro-Wilk test was applied to examine if the variables were normally distributed. For primary analyses, group differences in metabolite levels between controls and deletion carriers, and between deletion carriers with or without psychotic symptoms were tested using independent-sample t-tests (alpha level: 0.05). Effect sizes were estimated with Cohen’s d. Linear regression was used to test the age and age-by-neurometabolite interactions effects in deletion carriers *vs* controls. For secondary analyses, multiple linear regression was employed to predict neurometabolite levels in the hippocampus in the deletion carrier subgroup based on other neuroimaging variables such as the percentage of hippocampal atrophy and hippocampal BOLD variability. Finally, linear regression was used to test age effects and age-by-metabolite interactions in the hippocampus of deletion carriers with or without psychosis symptoms. The Benjamini-Hochberg procedure controlling for false discovery rates (FDR) was employed to adjust for multiple comparisons (62). All the reported p-values are FDR-corrected.

## Results

### Participants

One hundred and four participants, including 45 controls and 60 deletion carriers successfully underwent MRI and MRS acquisition. Controls and deletion carriers were matched for age and sex (Table 1).

Quality control performed as described above resulted in the exclusions of 2 participants for the ACC, 12 for the hippocampus and 9 for the STC. The remaining data yielded low fitting errors respectively for Glx H_2_O and GABA+H_2_O (ACC: 4.74 + 1.36/7.76+2.18; hippocampus; 8.15+4.74/9.84+4.3; STC:6.54+ 5.5/8+3.3).

### Metabolites Concentration: Controls vs Deletion Carriers

Significantly increased Glx levels were found in deletion carriers compared to controls in the hippocampus (t90 = 4.7, p < 0.001, d = 1.1) and STG (t93 = 2.9, p = 0.011, d = 0.6), but not in ACC (t100 = -0.38, p = 0.69, d = -0.08). Significant decreases in GABA+ levels were found in deletion carriers vs controls in the hippocampus (t90 = 6.6, p < 0.001, d = -1.5) but not in STG (t93 = -1, p = 0.6, d = -0.24) or ACC (t100 = -0.66, p = 0.51, d = -0.13) (Fig.2).

**Figure 2.**
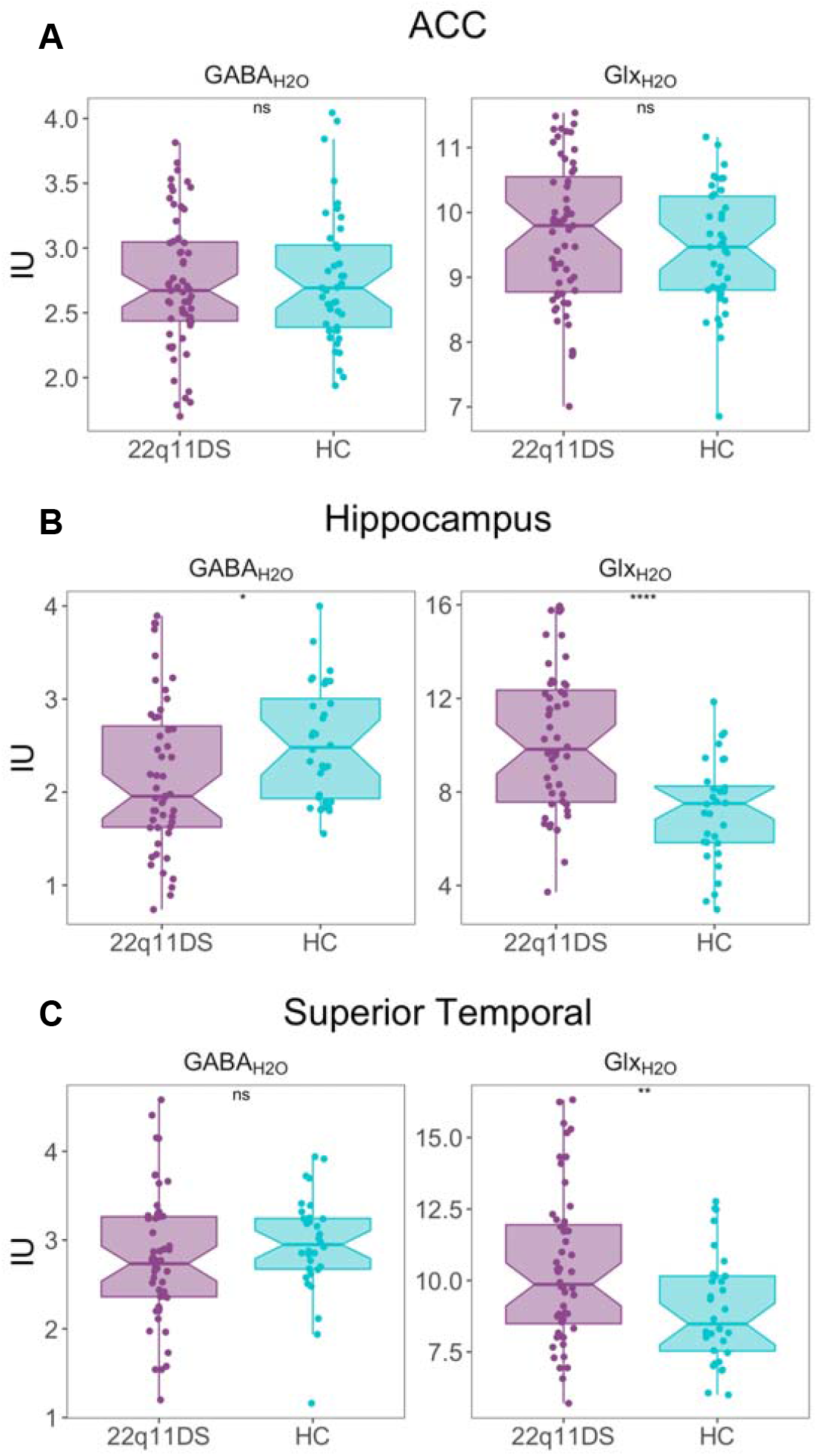
Comparison between controls and deletion carriers. GABA+ and Glx levels in: **a)** Anterior cingulate cortex (ACC), **b)** Hippocampus and **c)** Superior Temporal Cortex (STG). Compared to controls, deletion carriers were found to have higher levels of Glx in the hippocampus and STG and lower levels of GABA+ in the hippocampus. The lines in the boxplots indicate the median, the hinders of the boxplot correspond to the first and forth quartiles and the bars indicate 95% confidence interval. IU= international units; ns= non-significant; * p< .05, ** p< 0.001. Glx = glutamine + glutamate, GABA+ = GABA + macromolecules.

### Correlation between metabolites and age in Deletion carriers vs Controls

A simple linear regression was used to examine the role of age on neurometabolites in deletion carriers *vs* healthy controls. No significant age or age-by-group interaction effects were found for any of the ROIs (ACC, hippocampus, STC) and neurometabolites (Fig.1, Supplementary Materials).

### Metabolites Concentration in Deletion carriers: With vs Without Psychosis

Statistically significant increases were found between deletion carriers with or without psychotic symptoms for Glx concentration in the hippocampus (t50 = 3, p = 0.007, d = 0.91) but not in ACC (t55 = 0.18, p = 0.86, d = 0.05), or STC (t53 = -0.98, p = 0.92, d = -0.03). For GABA+, no differences were found (hippocampus (t50 = -0.42, p =0.67, d = -0.13), STC (t93 = 0.059, p = 0.96, d = 0.018), ACC (t53 = 0.17, p = 0.87, d = 0.05)) (fig.3).

**Figure 3.**
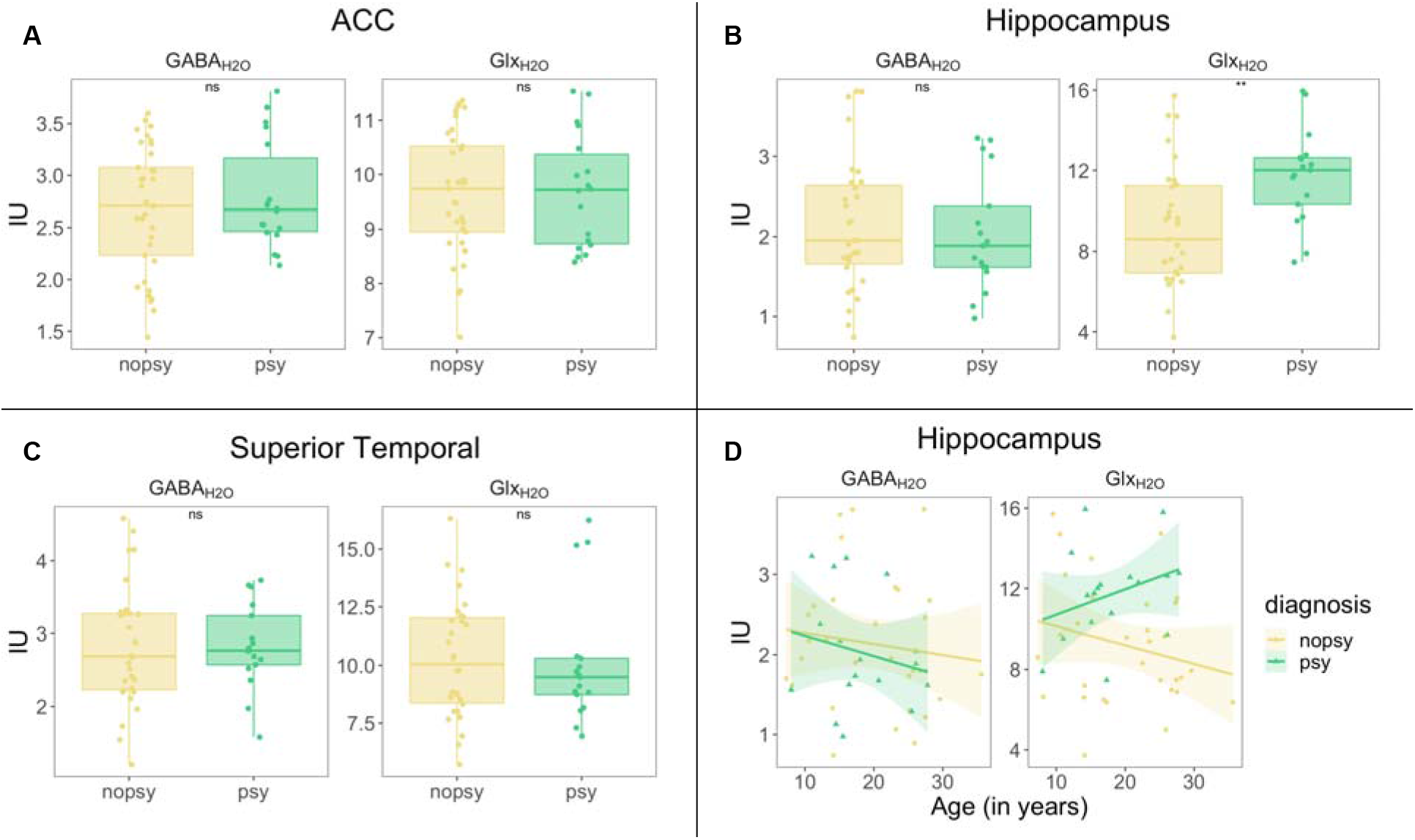
Comparison between deletion carriers with or without psychotic symptoms. GABA+ and Glx levls in **a)** Anterior cingulate cortex (ACC), **b)** Hippocampus and **c)** Superior Temporal Gyrus (STG). Deletion carriers with psychotic symptoms were found to have higher levels of Glx in the hippocampus compared to deletion carriers without psychotic symptoms. Subfigure **d)** indicates the correlation between hippocampal GABA+ and Glx and age in deletion carriers with or without psychotic symptoms. There was a significant age-by-group interaction for hippocampal Glx (R2= 0.22, F(3, 47) = 3.9, p = 0.014) but not for hippocampal GABA+ (R2= 0.03, F(3, 47) = 0.44, p = 0.75). The lines in the boxplots indicate the median, the hinders of the boxplot correspond to the first and forth quartiles and the bars indicate 95% confidence interval. IU= international units; ns= non-significant; * p< .05, ** p< 0.001. Glx = glutamine + glutamate, GABA+= GABA + macromolecules.

### Correlation between hippocampal metabolites and other brain measures in Deletion carriers

Given the increased levels of hippocampal Glx in deletion carriers, we investigated its relationship with neuroimaging measures indicative of hippocampal atrophy or activity in the deletion carrier subgroup. A multiple regression was run to predict hippocampal Glx based on the percentage of hippocampal atrophy and hippocampal BOLD variability. For this analysis, we included only data from deletion carriers with good ^1^H-MRS, rs-fMRI, and longitudinal T1-weighted data. Of the 53 deletion carriers included into the resting state fMRI analysis, 5 were excluded due to excessive movement defined as translation or rotation higher than 3 mm. Structural longitudinal data were available for all these participants. Both hippocampal atrophy (β_atrophy_= -0.39, p < 0.05) and BOLD variability (β_BOLD_= 0.4, p < 0.05) significantly predicted hippocampal Glx, leading to an overall significant model F(3, 45) = 10.2, p = 0.0003, R2 = 0.377 (fig.4).

**Figure 4:**
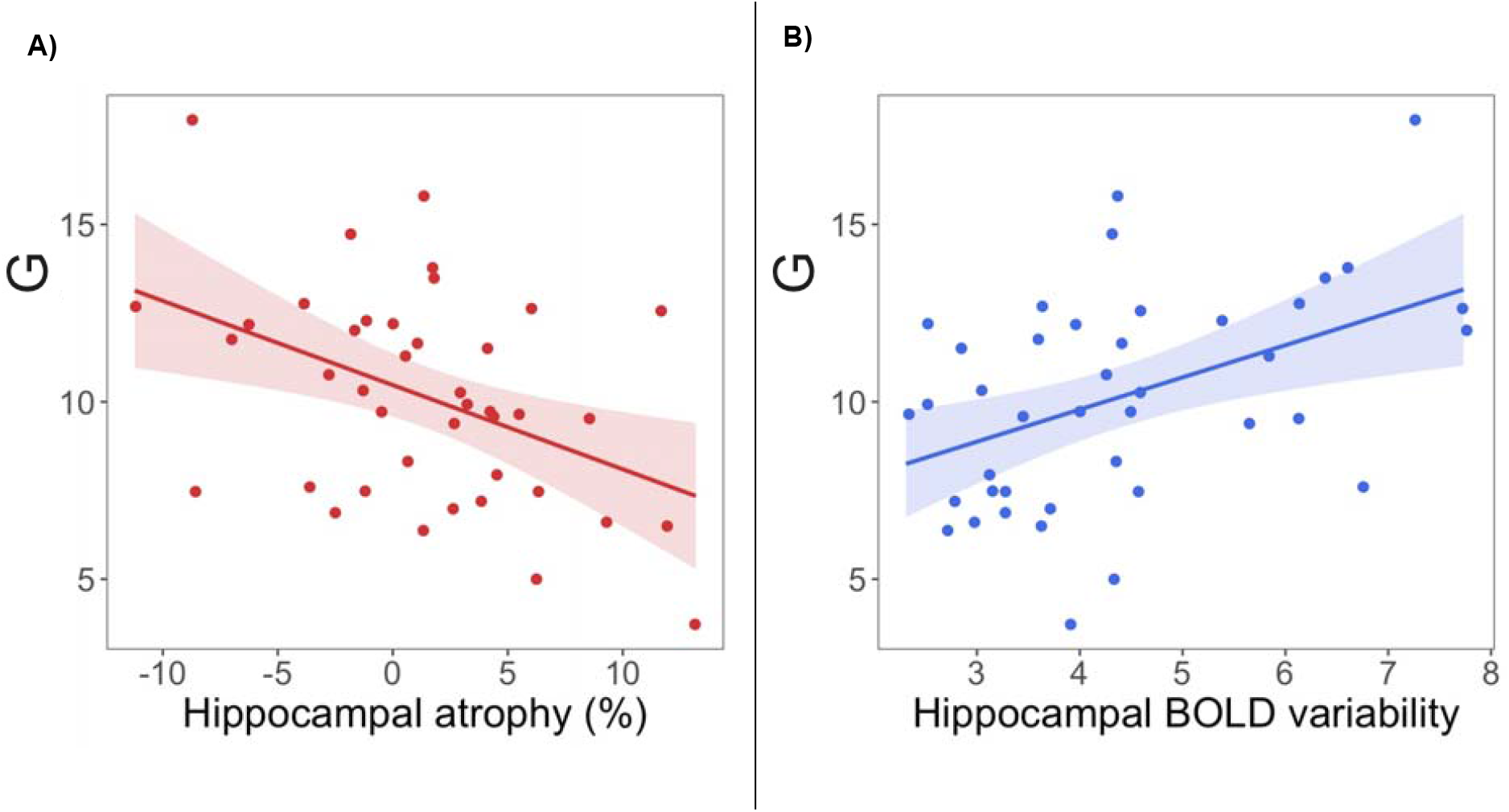
Correlation between hippocampal Glx and other neuroimaging variables in deletion carriers. In 22q11 deletion carriers, hippocampal Glx levels were a) negatively correlated with hippocampal atrophy, defined as the percentage of volume loss or increase with respect with the initial volume; and b) positively correlated with hippocampal BOLD variability.

### Correlation between hippocampal metabolites and age in Deletion carriers With vs Without Psychosis

A simple linear regression was used to examine the role of age on neurometabolites in deletion carriers with *vs* without psychotic symptoms. Results revealed significant age-by-group interaction for hippocampal Glx (R^2^= 0.22, F(3, 47) = 3.9, p = 0.014) whereby Glx levels increased with age in deletion carriers with psychotic symptoms and decreased with age in the non-symptomatic group. No significant age effects were found for hippocampal GABA+ (R^2^= 0.03, F(3, 47) = 0.44, p = 0.75).

## Discussion

Our results offer preliminary evidence for an alteration of the excitatory-inhibitory balance in temporo-limbic regions in 22q11 deletion carriers. Interestingly, some of the genes within the 22q11.2 deletion are implicated in glutamatergic and GABAergic neural transmission (63) and disrupted migration and placement of cortical interneurons and pyramidal cells (64–66). In particular, *PRODH* (proline dehydrogenase), a gene widely expressed in the brain, encodes for an enzyme that catalyzes the first step in the catabolism of proline to glutamate (67). Additionally, proline can modulate glutamatergic synaptic transmission either by directly activating glutamate receptors or acting as a co-agonist of NMDAR (68, 69). Thus, haploinsufficiency of *PRODH* may increase the levels of proline and affect glutamatergic transmission and glutamate concentration.

We identified increased levels of Glx in the STG and hippocampus and decreased levels of GABA in the hippocampus of deletion carriers compared to controls. By contrast, Glx and GABA levels were preserved in the ACC. Contrary to our expectations, we did not find any difference in the levels of Glx and GABA+ in the ACC of deletion carriers as compared to controls, nor in deletion carriers with psychotic symptoms compared to those without symptoms. However, this result is in line with previous studies showing a lack of differences of Glx levels in ACC in deletion carriers (19), suggesting that glutamatergic and GABAergic dysfunctions in the ACC may not be directly associated with the chromosome 22q11 deletion, nor an early emerging feature in deletion carriers presenting psychotic symptoms. Similarly, in previous research there was no evidence for glutamatergic dysregulation in the thalamus or the basal ganglia of deletion carriers (19) Overall, these findings indicate that glutamatergic dysregulation differentially affects distinct brain regions in 22q11DS, converging towards a model characterized by early dysfunction in temporo-limbic brain regions.

Enhanced glutamatergic signaling in the hippocampus was the only feature that characterized deletion carriers with emerging psychosis. Hippocampal vulnerability for the effects of glutamate may be mediated by differences between the neocortex and hippocampus at the cytoarchitectonic and neuronal level. Interestingly, in the hippocampus pyramidal layers are densely packed with glutamatergic cells, which account for over 90% of the overall number of neurons (70). Moreover, hippocampal glutamatergic neurons have unique cellular properties including higher excitability (lower firing threshold) (71) to optimize hippocampal functions such as learning and memory (72). Collectively, these features imply that the hippocampus is potentially more vulnerable to regional glutamatergic pathology and shifts in the excitatory/inhibitory balance than other brain regions (72). The identified hippocampal glutamatergic dysregulation may be the driver of previous volumetric and functional abnormalities identified in 22q11DS (41). Specifically, studies have shown that deletion carriers experiencing psychotic symptoms displayed a pattern of decreased connectivity with prefrontal cortex and increased connectivity with basal ganglia (43) and underwent progressive hippocampal atrophy during late adolescence (42). We therefore examined the potential role of hippocampal atrophy and BOLD signal variability on glutamatergic levels.

Notably, we found that hippocampal glutamate is positively associated with BOLD variability, which reflects moment-to-moment variability in brain activity as measured by resting-state fMRI (73–75). Previous studies have shown a correlation between fMRI measured connectivity – both during tasks and rest – and glutamate within the same brain area but also with distant regions (76). High BOLD variability has been related to poorer cognitive functions, as measured by a working memory n-back task (77), although other studies have shown an increase of BOLD variability with age in some brain regions, but not the hippocampus (52). A potential explanation is that increased glutamatergic activity and BOLD variability may be detrimental for cognition in deletion carriers with psychotic symptoms.

We additionally found that Glx levels were negatively associated with hippocampal atrophy in 22q11 deletion carriers. While the present findings do not imply causality, there is evidence that an early glutamatergic dysregulation and metabolic increase in the CA1 region of the hippocampus is subsequently followed by regional hippocampal atrophy, under the hypothesis that excessive elevation of glutamate may cause cells death via excitotoxicity, as proven by the administration of glutamate antagonists to prevent neuronal loss (78, 79). Excessive glutamate levels have furthermore been involved in psychosis, as studies in patients with chronic schizophrenia have shown a negative correlation between glutamate levels and hippocampal volume (45). Early models of psychosis further support the existence of a cascade involving early glutamatergic dysregulation leading to volume loss, as it has been shown that although glutamatergic hyperactivity in the hippocampus is present as early as the CHR status, only focal hippocampal atrophy predicted the transition to a full-blown psychotic disorder (17). Overall, the observed findings are in line with current pathophysiological models of psychosis positing that the earliest triggering factor may be the hypofunction of NMDAR on PVI, leading to enhanced glutamatergic neural transmission in the hippocampus that would cause augmented dopaminergic release in the striatum and over time hippocampal volume loss (1).

Finally, we identified divergent developmental trajectories of Glx levels in deletion carriers with or without psychotic symptoms, where psychotic individuals showed an increasing Glx trajectory, contrasting with progressively decreasing Glx levels in non-psychotic individuals. These findings, although preliminary, mirror longitudinal evidence for hippocampal atrophy and hypoconnectivity with prefrontal regions at the transition between adolescence and adulthood (42, 43). However, due to the inherent limitations of the MRS technique to distinguish between intra- and extracellular compartments, the interpretation of an increase of Glx from childhood to adulthood is not straightforward. Previous studies have shown that there is a decrease of glutamate levels in the prefrontal cortex of healthy individuals during early adulthood (80). Typically, during postnatal development synaptic pruning of glutamatergic synapses is necessary for physiological brain development (81). Impairment of adolescent pruning and synaptic remodeling in the hippocampus can lead to the disruption of learning processes during adulthood (82), as observed in animal models of schizophrenia and autism spectrum disorder (83). While speculative, one possible explanation for the observed lack of Glx decrease with age may be a dysregulation in the pruning of glutamatergic synapses in deletion carriers who develop psychotic symptoms. Further longitudinal studies are needed to confirm this initial finding.

### Limitations

One main conceptual limitation of the present study is that MRS at 3T does not allow the distinction between glutamate and glutamine (84). Future studies using higher magnetic field strength should further examine glutamate concentration in deletion carriers. Another limitation is that due to the later starting date of MRS data collection, longitudinal data was available only for the structural MRI data but not MRS data, which was cross-sectional. Thus, although our results are suggestive of a relationship between hippocampal atrophy and glutamate levels, we cannot infer the temporal directionality of this association.

## Conclusions

Our results support the idea that glutamatergic dysfunction in 22q11DS is region-specific, in that it primarily affects temporo-limbic brain structures. Additionally, evidence of further enhanced glutamatergic tone in the hippocampus of subjects experiencing psychotic symptoms point to a relationship between glutamatergic dysregulation and disease progression. Notably, the observed findings corroborate neurobiological theories of schizophrenia considering the hippocampus as the first area affected in early psychosis and the driver of downstream dopaminergic hyperactivity (1, 79). Overall, glutamatergic dysfunction in the hippocampus might represent a promising target for early intervention.

## Data Availability

All data produced in the present study are available upon reasonable request to the authors

## Acknowledgements and Disclosures

This study was funded by research grants from the Swiss National Science Foundation (grants 324730-144260 and 320030-179404 to Dr. Eliez) and a National Center of Competence in Research Synapsy grant (grant 51NF40-185897 to Dr. Eliez).

The authors thank all the families who contributed to the study as well as the family associations (Generation 22, Connect 22, and Relais 22) for their ongoing support. The authors also thank Virginie Pouillard, Eva Micol, and Tereza Kotalova for coordinating the project; the managers and operators of the MRI platforms at Campus Biotech Roberto Martuzzi, Loan Mattera, and Olivier Reynaud; and Karin Bortolin for her help in the acquisition of the data. Finally, the authors thank Sinyeob Ahn from Siemens for proofreading the manuscript.

## References

1. Lieberman JA, First MB (2018): Psychotic Disorders. (A. H. Ropper, editor) N Engl J Med. 379: 270–280.

2. Olney JW, Farber N (1995): Glutamate Receptor Dysfunction and Schizophrenia. Arch Gen Psychiatry. 52: 998.

3. Lisman JE, Coyle JT, Green RW, Javitt DC, Benes FM, Heckers S, Grace AA (2008): Circuit-based framework for understanding neurotransmitter and risk gene interactions in schizophrenia. Trends Neurosci. 31: 234–242.

4. Beneyto M, Kristiansen L V, Oni-Orisan A, McCullumsmith RE, Meador-Woodruff JH (2007): Abnormal Glutamate Receptor Expression in the Medial Temporal Lobe in Schizophrenia and Mood Disorders. Neuropsychopharmacology. 32: 1888–1902.

5. Javitt D, Zukin S (1991): Recent advances in the phencyclidine model of schizophrenia. Am J Psychiatry. 148: 1301–1308.

6. Malhotra A (1996): NMDA Receptor Function and Human Cognition: The Effects of Ketamine in Healthy Volunteers. Neuropsychopharmacology. 14: 301–307.

7. Pollak TA, Lennox BR, Müller S, Benros ME, Prüss H, Tebartz van Elst L, et al. (2020): Autoimmune psychosis: an international consensus on an approach to the diagnosis and management of psychosis of suspected autoimmune origin. The Lancet Psychiatry. 7: 93–108.

8. Steiner J, Walter M, Glanz W, Sarnyai Z, Bernstein HG, Vielhaber S, et al. (2013): Increased prevalence of diverse N-methyl-D-aspartate glutamate receptor antibodies in patients with an initial diagnosis of schizophrenia: Specific relevance of IgG NR1a antibodies for distinction from N-methyl-D-aspartate glutamate receptor encephalitis. JAMA Psychiatry. 70: 271–278.

9. Ripke S, Neale B, Corvin A (2014): Biological insights from 108 schizophrenia-associated genetic loci. Nature. 511: 421–427.

10. Merritt K, Egerton A, Kempton MJ, Taylor MJ, McGuire PK (2016): Nature of glutamate alterations in schizophrenia a meta-analysis of proton magnetic resonance spectroscopy studies. JAMA Psychiatry. 73: 665–674.

11. Merritt K, McGuire PK, Egerton A, Aleman A, Block W, Bloemen OJN, et al. (2021): Association of Age, Antipsychotic Medication, and Symptom Severity in Schizophrenia With Proton Magnetic Resonance Spectroscopy Brain Glutamate Level. JAMA Psychiatry. 78: 667.

12. Thakkar KN, Rösler L, Wijnen JP, Boer VO, Klomp DWJ, Cahn W, et al. (2017): 7T Proton Magnetic Resonance Spectroscopy of Gamma-Aminobutyric Acid, Glutamate, and Glutamine Reveals Altered Concentrations in Patients With Schizophrenia and Healthy Siblings. Biol Psychiatry. 81: 525–535.

13. Wenneberg C, Nordentoft M, Rostrup E, Glenthøj LB, Bojesen KB, Fagerlund B, et al. (2020): Cerebral Glutamate and Gamma-Aminobutyric Acid Levels in Individuals at Ultra-high Risk for Psychosis and the Association With Clinical Symptoms and Cognition. Biol Psychiatry Cogn Neurosci Neuroimaging. 5: 569–579.

14. Marenco S, Meyer C, Kuo S, Van Der Veen JW, Shen J, DeJong K, et al. (2016): Prefrontal GABA levels measured with magnetic resonance spectroscopy in patients with psychosis and unaffected siblings. Am J Psychiatry. 173: 527–534.

15. Reddy-Thootkur M, Kraguljac NV, Lahti AC (2020): The role of glutamate and GABA in cognitive dysfunction in schizophrenia and mood disorders – A systematic review of magnetic resonance spectroscopy studies. Schizophr Res.. doi: 10.1016/j.schres.2020.02.001.

16. Bossong MG, Antoniades M, Azis M, Samson C, Quinn B, Bonoldi I, et al. (2019): Association of Hippocampal Glutamate Levels with Adverse Outcomes in Individuals at Clinical High Risk for Psychosis. JAMA Psychiatry. 76: 199–207.

17. Provenzano FA, Guo J, Wall MM, Feng X, Sigmon HC, Brucato G, et al. (2020): Hippocampal Pathology in Clinical High-Risk Patients and the Onset of Schizophrenia. Biol Psychiatry. 87: 234–242.

18. Wang AM, Pradhan S, Coughlin JM, Trivedi A, Dubois SL, Crawford JL, et al. (2019): Assessing Brain Metabolism with 7-T Proton Magnetic Resonance Spectroscopy in Patients with First-Episode Psychosis. JAMA Psychiatry. 76: 314–323.

19. Rogdaki M, Hathway P, Gudbrandsen M, McCutcheon RA, Jauhar S, Daly E, Howes O (2019): Glutamatergic function in a genetic high-risk group for psychosis: A proton magnetic resonance spectroscopy study in individuals with 22q11.2 deletion. Eur Neuropsychopharmacol. 29: 1333–1342.

20. da Silva Alves F, Boot E, Schmitz N, Nederveen A, Vorstman J, Lavini C, et al. (2011): Proton magnetic resonance spectroscopy in 22q11 deletion syndrome. PLoS One. 6. doi: 10.1371/journal.pone.0021685.

21. Vingerhoets C, Tse DHY, van Oudenaren M, Hernaus D, van Duin E, Zinkstok J, et al. (2020): Glutamatergic and GABAergic reactivity and cognition in 22q11.2 deletion syndrome and healthy volunteers: A randomized double-blind 7-Tesla pharmacological MRS study. J Psychopharmacol. 34: 856–863.

22. Caballero A, Tseng KY (2016): GABAergic Function as a Limiting Factor for Prefrontal Maturation during Adolescence. Trends Neurosci. 39: 441–448.

23. Caballero A, Granberg R, Tseng KY (2016): Mechanisms contributing to prefrontal cortex maturation during adolescence. Neurosci Biobehav Rev. 70: 4–12.

24. Tseng KY, O’Donnell P (2007): Dopamine modulation of prefrontal cortical interneurons changes during adolescence. Cereb Cortex. 17: 1235–1240.

25. Larsen B, Cui Z, Adebimpe A, Pines A, Alexander-Bloch A, Bertolero M, et al. (2022): A developmental reduction of the excitation:inhibition ratio in association cortex during adolescence. Sci Adv. 8. doi: 10.1126/sciadv.abj8750.

26. Schneider Maude, Debbanè Martin, Bassett Anne S. CEW (2015): Psychiatric Disorders From Childhood to Adulthood in 22q11.2 Deletion Syndrome: Results From the International Consortium on Brain and Behavior in 22q11.2 Deletion Syndrome. Am J Psychiatry. 171: 627–639.

27. Meechan DW, Maynard TM, Fernandez A, Karpinski BA, Rothblat LA, LaMantia AS (2015): Modeling a model: Mouse genetics, 22q11.2 Deletion Syndrome, and disorders of cortical circuit development.pdf. Prog Neurobiol. 130: 1–28.

28. Meechan DW, Tucker ES, Maynard TM, LaMantia AS (2009): Diminished dosage of 22q11 genes disrupts neurogenesis and cortical development in a mouse model of 22q11 deletion/DiGeorge syndrome. Proc Natl Acad Sci U S A. 106: 16434–16445.

29. Earls LR, Bayazitov IT, Fricke RG, Berry RB, Illingworth E, Mittleman G, Zakharenko SS (2010): Dysregulation of presynaptic calcium and synaptic plasticity in a mouse model of 22q11 deletion syndrome. J Neurosci. 30: 15843–15855.

30. Mukherjee A, Carvalho F, Eliez S, Caroni P, Mukherjee A, Carvalho F, et al. (2019): Long-Lasting Rescue of Network and Cognitive Dysfunction in a Genetic Schizophrenia Model Article Long-Lasting Rescue of Network and Cognitive Dysfunction. Cell. 1–16.

31. Schaer M, Debbané M, Bach Cuadra M, Ottet MC, Glaser B, Thiran JP, Eliez S (2009): Deviant trajectories of cortical maturation in 22q11.2 deletion syndrome (22q11DS): A cross-sectional and longitudinal study. Schizophr Res. 115: 182–190.

32. Padula MC, Scariati E, Schaer M, Eliez S (2018): A mini review on the contribution of the anterior cingulate cortex in the risk of psychosis in 22q11.2 deletion syndrome. Front Psychiatry. 9: 9–14.

33. Zöller D, Padula MC, Sandini C, Schneider M, Scariati E, Van De Ville D, et al. (2018): Psychotic symptoms influence the development of anterior cingulate BOLD variability in 22q11.2 deletion syndrome. Schizophr Res. 193: 319–328.

34. Schmitt JE, Vandekar S, Yi J, Calkins ME, Ruparel K, Roalf DR, et al. (2015): Archival Report Aberrant Cortical Morphometry in the 22q11. 2 Deletion Syndrome. Biol Psychiatry. 78: 135–143.

35. Chow EWC, Ho A, Wei C, Voormolen EHJ, Crawley AP, Bassett AS (2011): Association of Schizophrenia in 22q11.2 Deletion Syndrome and Gray Matter Volumetric Deficits in the Superior Temporal Gyrus. Am J Psychiatry. 168: 522–529.

36. Sun D, Ching CRK, Lin A, Forsyth JK, Kushan L, Vajdi A, et al. (2017): Large-scale mapping of cortical alterations in 22q11. 2 deletion syndromeLJ: Convergence with idiopathic psychosis and effects of deletion size. Mol Psychiatry.. doi: 10.1038/s41380-018-0078-5.

37. Bagautdinova J, Zöller D, Schaer M, Padula MC, Mancini V, Schneider M, Eliez S (2021): Altered cortical thickness development in 22q11.2 deletion syndrome and association with psychotic symptoms. Mol Psychiatry.. doi: 10.1038/s41380-021-01209-8.

38. Cantonas LM, Mancini V, Rihs TA, Rochas V, Schneider M, Eliez S, Michel CM (2021): Abnormal Auditory Processing and Underlying Structural Changes in 22q11.2 Deletion Syndrome. Schizophr Bull. 47: 189–196.

39. Mancini V, Rochas V, Seeber M, Roehri N, Rihs TA, Ferat V, et al. (2022): Aberrant Developmental Patterns of Gamma-Band Response and Long-Range Communication Disruption in Youths With 22q11.2 Deletion Syndrome. Am J Psychiatry. 179: 204–215.

40. Mancini V, Zöller D, Schneider M, Schaer M, Eliez S (2020): Abnormal Development and Dysconnectivity of Distinct Thalamic Nuclei in Patients With 22q11.2 Deletion Syndrome Experiencing Auditory Hallucinations. Biol Psychiatry Cogn Neurosci Neuroimaging. 5: 875–890.

41. Rogdaki M, Gudbrandsen M, Mccutcheon RA, Blackmore CE, Brugger S, Ecker C, et al. (2020): Magnitude and heterogeneity of brain structural abnormalities in 22q11. 2 deletion syndromeLJ: a meta-analysis. Mol Psychiatry.. doi: 10.1038/s41380-019-0638-3.

42. Mancini V, Sandini C, Padula MC, Zöller D, Schneider M, Schaer M, Eliez S (2020): Positive psychotic symptoms are associated with divergent developmental trajectories of hippocampal volume during late adolescence in patients with 22q11DS. Mol Psychiatry. 25: 2844–2859.

43. Delavari F, Sandini C, Zöller D, Mancini V, Bortolin K, Schneider M, et al. (2020): Dysmaturation observed as altered hippocampal functional connectivity at rest is associated with the emergence of positive psychotic symptoms in patients with 22q11 deletion syndrome. Biol Psychiatry.. doi: 10.1016/j.biopsych.2020.12.033.

44. Alver M, Mancini V, Läll K, Schneider M, Romano L, Estonian Biobank Research Team, et al. (2022): Contribution of schizophrenia polygenic burden to longitudinal phenotypic variance in 22q11.2 deletion syndrome. Mol Psychiatry. 1–10.

45. Kraguljac N V., White DM, Reid MA, Lahti AC (2013): Increased hippocampal glutamate and volumetric deficits in unmedicated patients with schizophrenia. JAMA Psychiatry. 70: 1294–1302.

46. Miller TJ, Mcqlashan TH, Rosen JL, Cadenhead K, Ventura J, Mcfarlane W, et al. (1995): Prodromal Assessment With the Structured Interview for Prodromal Syndromes and the Scale of Prodromal SymptomsLJ: Predictive Validity, Interrater Reliability, and Training to Reliability. Schizophr Bull. 703–716.

47. Mescher M, Merkle H, Kirsch J, Garwood M, Gruetter R (1998): Simultaneousin vivo spectral editing and water suppression. NMR Biomed. 11: 266–272.

48. Saleh MG, Rimbault D, Mikkelsen M, Oeltzschner G, Wang AM, Jiang D, et al. (2019): Multi-vendor standardized sequence for edited magnetic resonance spectroscopy. Neuroimage. 189: 425–431.

49. Fischl B (2012): FreeSurfer. Neuroimage. 62: 774–781.

50. Desikan RS, Se F, Fischl B, Quinn BT, Dickerson BC, Blacker D, et al. (2006): An automated labeling system for subdividing the human cerebral cortex on MRI scans into gyral based regions of interest. 31: 968–980.

51. Iglesias JE, Augustinack JC, Nguyen K, Player CM, Player A, Wright M, et al. (2015): A computational atlas of the hippocampal formation using ex vivo, ultra-high resolution MRI: Application to adaptive segmentation of in vivo MRI. Neuroimage. 115. doi: 10.1016/j.neuroimage.2015.04.042.

52. Zöller D, Schaer M, Scariati E, Padula MC, Eliez S, Van De Ville D (2017): Disentangling resting-state BOLD variability and PCC functional connectivity in 22q11.2 deletion syndrome. Neuroimage. 149. doi: 10.1016/j.neuroimage.2017.01.064.

53. Chao-gan Y, Yu-feng Z (2010): DPARSFLJ: a MATLAB toolbox for “ pipeline “ data analysis of resting-state fMRI. 4: 1–7.

54. Alemán-Gómez Y, Melie-García L, Valdés-Hernandez P (2006): IBASPM: Toolbox for automatic parcellation of brain structures. resented 12th Annu Meet Organ Hum Brain Mapp..

55. Ashburner J, Friston KJ (2005): Unified segmentation. Neuroimage. 26: 839–851.

56. Ashburner J (2007): A fast diffeomorphic image registration algorithm. Neuroimage. 38: 95–113.

57. Power JD, Barnes KA, Snyder AZ, Schlaggar BL, Petersen SE (2012): Spurious but systematic correlations in functional connectivity MRI networks arise from subject motion. Neuroimage. 59: 2142–2154.

58. Tzourio-Mazoyer N, Landeau B, Papathanassiou D, Crivello F, Etard O, Delcroix N, et al. (2002): Automated Anatomical Labeling of Activations in SPM Using a Macroscopic Anatomical Parcellation of the MNI MRI Single-Subject Brain. Neuroimage. 15: 273–289.

59. Joined PL, Herald TM, Globe TB, Guardian T (2010): Gannet: A Batch-Processing Tool for theQuantitative Analysis of Gamma-AminobutyricAcid–Edited MR Spectroscopy Spectra. Foundations. 1–13.

60. Gasparovic C, Song T, Devier D, Bockholt HJ, Caprihan A, Mullins PG, et al. (2006): Use of tissue water as a concentration reference for proton spectroscopic imaging. Magn Reson Med. 55: 1219–1226.

61. Saleh MG, Papantoni A, Mikkelsen M, Hui SCN, Oeltzschner G, Puts NA, et al. (2020): Effect of Age on GABA+ and Glutathione in a Pediatric Sample. Am J Neuroradiol. 41: 1099–1104.

62. Benjamini Y, Hochberg Y (1995): Controlling the False Discovery Rate: A Practical and Powerful Approach to Multiple Testing. R Stat Soc. 57: 289–300.

63. Motahari Z, Moody SA, Maynard TM, Lamantia AS (2019): In the line-up: Deleted genes associated with DiGeorge/22q11.2 deletion syndrome: Are they all suspects? J Neurodev Disord. 11: 1–28.

64. Toritsuka M, Kimoto S, Muraki K, Landek-salgado MA, Yoshida A (2013): Deficits in microRNA-mediated Cxcr4/Cxcl12 signaling in neurodevelopmental deficits in a 22q11 deletion syndrome mouse model. Proc Natl Acad Sci. 110: 4–9.

65. Meechan DW, Tucker ES, Maynard TM, LaMantia AS (2012): Cxcr4 regulation of interneuron migration is disrupted in 22q11.2 deletion syndrome. Proc Natl Acad Sci U S A. 109: 18601–18606.

66. Molinard-Chenu A, Dayer A (2018): The Candidate Schizophrenia Risk Gene DGCR2 Regulates Early Steps of Corticogenesis. Biol Psychiatry. 83: 692–706.

67. Mitsubuchi H, Nakamura K, Matsumoto S, Endo F (2014): Biochemical and clinical features of hereditary hyperprolinemia. Pediatr Int. 56: 492–496.

68. Cohen SM, Nadler JV (1997): Proline-induced potentiation of glutamate transmission. Brain Res. 761: 271–282.

69. Cohen SM, Nadler JV (1997): Proline-induced inhibition of glutamate release in hippocampal area CA1. Brain Res. 769: 333–339.

70. Olbrich H-G, Braak H (1985): Ratio of pyramidal cells versus non-pyramidal cells in sector CA1 of the human Ammon’s horn. Anat Embryol (Berl). 173: 105–110.

71. Marín-Burgin A, Mongiat LA, Pardi MB, Schinder AF (2012): Unique Processing During a Period of High Excitation/Inhibition Balance in Adult-Born Neurons. Science (80-). 335: 1238–1242.

72. Tamminga CA, Stan AD, Wagner AD (2010): The Hippocampal Formation in Schizophrenia. Am J Psychiatry. 167: 1178–1193.

73. Garrett DD, Samanez-Larkin GR, MacDonald SWS, Lindenberger U, McIntosh AR, Grady CL (2013): Moment-to-moment brain signal variability: A next frontier in human brain mapping? Neurosci Biobehav Rev. 37: 610–624.

74. Garrett DD, Kovacevic N, McIntosh AR, Grady CL (2013): The Modulation of BOLD Variability between Cognitive States Varies by Age and Processing Speed. Cereb Cortex. 23: 684–693.

75. Garrett DD, Kovacevic N, McIntosh AR, Grady CL (2010): Blood Oxygen Level-Dependent Signal Variability Is More than Just Noise. J Neurosci. 30: 4914–4921.

76. Duncan NW, Wiebking C, Northoff G (2014): Associations of regional GABA and glutamate with intrinsic and extrinsic neural activity in humans-A review of multimodal imaging studies. Neurosci Biobehav Rev. 47: 36–52.

77. Boylan MA, Foster CM, Pongpipat EE, Webb CE, Rodrigue KM, Kennedy KM (2021): Greater BOLD Variability is Associated With Poorer Cognitive Function in an Adult Lifespan Sample. Cereb Cortex. 31: 562–574.

78. Schobel SA, Chaudhury NH, Khan UA, Paniagua B, Styner MA, Asllani I, et al. (2013): Imaging Patients with Psychosis and a Mouse Model Establishes a Spreading Pattern of Hippocampal Dysfunction and Implicates Glutamate as a Driver. Neuron. 78: 81–93.

79. Lieberman JA, Girgis RR, Brucato G, Moore H, Provenzano F, Kegeles L, et al. (2018): Hippocampal dysfunction in the pathophysiology of schizophrenia: a selective review and hypothesis for early detection and intervention. Mol Psychiatry. 23: 1764–1772.

80. Marsman A, Mandl RCW, van den Heuvel MP, Boer VO, Wijnen JP, Klomp DWJ, et al. (2013): Glutamate changes in healthy young adulthood. Eur Neuropsychopharmacol. 23: 1484–1490.

81. Paolicelli RC, Bolasco G, Pagani F, Maggi L, Scianni M, Panzanelli P, et al. (2011): Synaptic Pruning by Microglia Is Necessary for Normal Brain Development. Science (80-). 333: 1456–1458.

82. Afroz S, Parato J, Shen H, Smith SS (2016): Synaptic pruning in the female hippocampus is triggered at puberty by extrasynaptic GABAA receptors on dendritic spines. Elife. 5: 1–23.

83. van Spronsen M, Hoogenraad CC (2010): Synapse Pathology in Psychiatric and Neurologic Disease. Curr Neurol Neurosci Rep. 10: 207–214.

84. Hancu I (2009): Optimized glutamate detection at 3T. J Magn Reson Imaging. 30: 1155–1162.

