## Supplementary figure 1 for "Excitatory inhibitory imbalance underlies hippocampal atrophy in individuals with 22q11.2 Deletion Syndrome with psychotic symptoms"

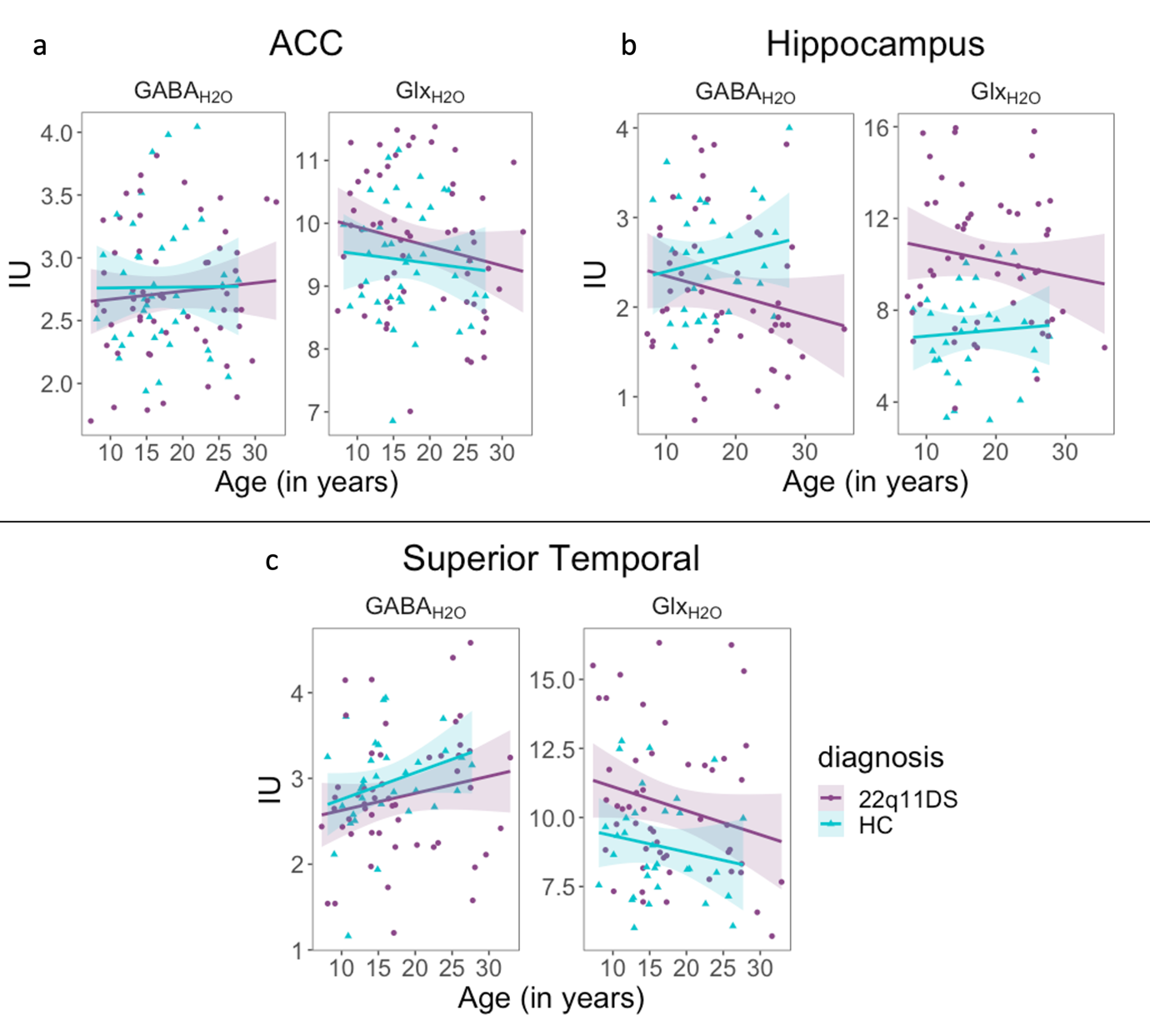

**Supplementary figure 1 Developmental trajectories of Glx And GABA+ in deletion carriers and controls**

GABA+ and Glx developmental trajectories in **a)** Anterior cingulate cortex (ACC), **b)** Hippocampus and **c)** Superior Temporal Gyrus (STG). There was no significant age-by-group interaction for any of the ROIs and neurometabolite. IU= international units; Glx = glutamine + glutamate, GABA+= GABA + macromolecules.
